# Mapping Human Development Index and Nutritional Status: Identifying Factors Associated with Malnutrition in Indonesia

**DOI:** 10.64898/2026.06.24.26355946

**Authors:** Fhadilla Amelia, Arina Nurul Ihsani, Yoerdy Agusmal Saputra, Rohana Uly Pradita Siregar

## Abstract

**Background:** Indonesia, as an upper-middle-income country, is undergoing a nutrition transition. The persistent problem of undernutrition is now accompanied by rising overnutrition, creating a double burden of malnutrition. The Human Development Index (HDI) that measures socio-economic development, has shown steady improvements in Indonesia, yet nutritional disparities remain across provinces with varying HDI levels.

**Objective/Purpose:** This study aims to map the relationship between HDI and nutritional status by analyzing the prevalence of stunting, wasting, underweight, and overweight across different HDI categories and provinces in Indonesia. Additionally, this study explores the determinants of malnutrition to provide a deeper understanding of the factors influencing child nutrition status.

**Methods:** This ecological study used 2023 provincial-level data from the Indonesian Health Survey (SKI) and the Human Development Index (HDI) published by the Central Bureau of Statistics. Nutritional indicators (stunting, underweight, wasting, and overweight) were analyzed in relation to HDI and other determinants. Spearman’s correlation and Mann-Whitney U tests were used for statistical analysis. Spatial patterns were visualized through GIS mapping to explore geographic relationships between HDI levels and nutritional status.

**Results:** Correlational analysis showed that Medium HDI provinces had significantly higher rates of undernutrition than High-Very High HDI provinces and were significantly correlated with low birth weight, exclusive breastfeeding, proper IYCF, adequate micronutrient consumption, proper handwashing, proper sanitation, complete basic immunization, complete compulsory education, and HDI.

**Conclusion:** This study highlights the complex relationship between HDI and nutritional status, emphasizing the need for region-specific interventions. While improving HDI can help to reduce undernutrition, rising overweight prevalence requires targeted public health strategies. These findings offer valuable insights for policymakers to design holistic, multi-dimensional approaches to combat malnutrition in Indonesia.

## Background

Over the past decades, the Human Development Index (HDI) has shown a global upward trend which reflects substantial progress in health, education, and income (1). In line with this, Indonesia has shown a steady improvement in its HDI, increasing from 68.31 in 2013 to 74.39 in 2023, indicating notable socio-economic progress over the past decade (2,3). HDI is a composite measure developed by the United Nations Development Programme (UNDP) to assess and compare the overall socio-economic development of regions and countries (4). It encompasses three key dimensions: health, measured by life expectancy at birth; education, measured by the average years of schooling for adults aged 25 and older and the expected years of schooling for children; and standard of living, measured by Gross National Income (GNI) per capita adjusted for purchasing power parity (PPP) (5). As a multidimensional indicator, HDI offers a holistic view of development and its impact on health outcomes, including nutritional status.

Nutritional status reflects an individual’s health condition, primarily influenced by food consumption and the body’s ability to utilize nutrients. Key indicators regarding undernutrition include wasting, stunting, and underweight; while overnutrition is assessed through overweight and obesity(6). However, nutritional status extends beyond individual dietary intake and is shaped by multiple basic, underlying, and immediate determinants (7). Further, social determinants like income, education, unemployment, and access to healthcare also significantly influence malnutrition, contributing to 30–55% of overall health outcomes (8). Thus, understanding nutritional status within this broader framework is essential for addressing malnutrition effectively and developing targeted public health interventions.

The components of HDI are deeply interrelated with nutritional status. Education, particularly for women, plays a pivotal role in shaping dietary practices and healthcare utilization. Further, a study has demonstrated that higher levels of maternal education are linked to reduced rates of stunting and underweight in children (9). Income is another critical determinant as it influences a household’s capacity to afford food, food choices, clean water, sanitation facilities, and many decision makings that are necessary for maintaining optimum nutrition (10). Increased household income has been linked to reduced rates of underweight and food insecurity due to better access to available food resources (11). Access to healthcare, as reflected in the life expectancy in HDI, ensures that individuals can prevent and treat nutrition-related illnesses, including malnutrition (8). Moreover, higher life expectancy is often indicative of better nutritional status. Research has shown that adopting a healthier diet, shifting from unhealthy eating habits to those linked with longevity, is associated with an increase in life expectancy (12). These theoretical linkages and findings illustrate how HDI dimensions collectively shape the environment in which nutritional outcomes are determined.

Indonesia, as an upper-middle-income country, is experiencing a rapid nutrition transition. The persistent issues of undernutrition is being overshadowed by a growing prevalence of overnutrition (13). In 2023, the prevalence of stunting, wasting, underweight, and overweight among children under five were 21.5%, 8.5%, 15.9%, and 4.2%, respectively (14) . These statistics highlight a double burden of malnutrition in Indonesia, with undernutrition rates showing a persistent issue while overnutrition concerns are on the rise. On the other hand, Indonesia’s HDI has reached the high category. However, substantial variation remains across provinces, with HDI levels ranging from low to very high (2). This variation in HDI underscores the need to analyze how socio-economic development influences nutritional outcomes, making it crucial to explore the disparities in malnutrition across different HDI categories in Indonesia.

By mapping each province based on its HDI and examining the nutritional issues within these provinces presents a unique and valuable opportunity for understanding the nuanced relationship between development and nutrition. Despite the increasing awareness of the connection between socio-economic factors and nutritional outcomes, there is limited research exploring the direct impact of HDI on nutritional status in Indonesia. While previous studies have focused on specific components of HDI, such as income or education, few have analyzed the combined effect of HDI on nutritional status at the population level. This study addresses this gap by providing a comprehensive analysis of how HDI affects the prevalence of stunting, wasting, underweight, and overweight. By mapping these relationships, the study offers critical insights for policymakers to develop targeted interventions that address malnutrition through a multi-dimensional approach.

Thus, this study aims to present overlapping data on HDI and nutritional status through maps. It enables a clear visualization of how development status impacts nutritional outcomes across provinces with similar or differing HDI levels. Additionally, the study investigates the determinants of stunting, underweight, wasting, and overweight to provide a comprehensive understanding of factors correlating with nutritional status for children under five in Indonesia.

## Methods

### Study Design

This ecological study utilized aggregate data (percentages/prevalence) at the provincial level in Indonesia. The Human Development Index (HDI) data were sourced from the 2023 report by the Central Bureau of Statistics (BPS). Other data were obtained from the 2023 Indonesian Health Survey (SKI), conducted by the Ministry of Health. Indonesia’s 34 provinces served as the unit of analysis.

We used the conceptual framework of UNICEF (15), adapted to align with the availability of existing data in Indonesia. The immediate causes of malnutrition analyzed in this study included birth weight, breastfeeding, feeding practices, micronutrient intake, and immunization. The underlying causes considered were hand hygiene and sanitation, as well as parental education levels. Additionally, the Human Development Index (HDI) was included as a central focus of the analysis.

### Variables

The dependent variables in this study were the prevalence rates of malnutrition reported in the 2023 SKI. The independent variables included HDI, birth weight, breastfeeding, feeding practices, micronutrient intake, immunization, hand hygiene, sanitation, and parental education levels.

### Data Analysis

Descriptive statistics were used to summarize the characteristics of the data. Correlations between the independent and dependent variables were analyzed using Spearman’s rank correlation, as the dependent variable did not meet the normality assumption. Differences in malnutrition prevalence rates based on provincial HDI levels were examined using the Mann-Whitney U test. Provinces with an HDI <0.700 were categorized as having low or medium HDI, while provinces with an HDI ≥0.700 were classified as having high or very high HDI.

The relationship pattern between HDI and stunting, wasting, underweight, and overweight was observed using spatial analysis. First, HDI data for each province was added to the base map, and then the vector data of the independent variables were processed, and color symbols were chosen. Based on the data size, digital categories of high and low HDI variables were generated. Next, the dependent variable’s vector data was turned into a digital file by adding spatial data of malnutrition (such as stunting, wasting, underweight, or overweight) to the base map. The centroid symbol was then processed and chosen. Digital categories for large and small cases were created based on the malnutrition data.

The resulting color gradations and dot symbols did not represent ratios but instead showed ordinal numbers, such as high-low HDI and malnutrition prevalence categories. The color gradient reflected HDI levels, manually grouped into four classes: grey = very high, dark purple = high, light purple = medium, and red = low (5). Malnutrition (stunting, wasting, underweight, and overweight) was depicted using centroid markers that vary in size from large to small according to the prevalence of the corresponding nutritional status. Then, each marker was labeled with its respective percentage value. The size of the dot symbol was digitally created using simple markers or standardized symbols from QGIS software with stunting classification: very low to very high (16), wasting classification: very low to very high (16), underweight classification: no public health problem to severe (18), and overweight classification: very low to very high (16) with a dot size of 1 to 25. Finally, the spatially analyzed data were processed with a thematic overlay to reveal patterns of spatial relationships.

All analyses were conducted using SPSS version 28 and QGIS software version 3.38.3.

### Ethical Considerations

Ethical approval for the SKI survey was granted by the Ethics Committee of Poltekkes Jakarta II (LB.02.01.I/KE/L/287/2023). This study utilized aggregate data at provincial level. The data are publicly available and openly accessible which do not contain any personal or identifiable information. The research did not involve any direct interaction with human participants or access to confidential data. The author also confirmed to the authorized party that obtaining ethical approval for this analysis was not required. This use of secondary data ensures compliance with ethical guidelines for research while maintaining the privacy and anonymity of individuals.

## Results

### A HDI difference of the prevalence of stunting, underweight, wasting, and overweight in Indonesia

A spatial analysis was conducted by overlaying a map of nutritional status, including stunting, underweight, wasting, and overweight in children under five, and defined by Human Development Index (HDI). The color gradient signifies the category of HDI, whereas the dot symbol indicates each nutritional status prevalence.

**Fig 1.**
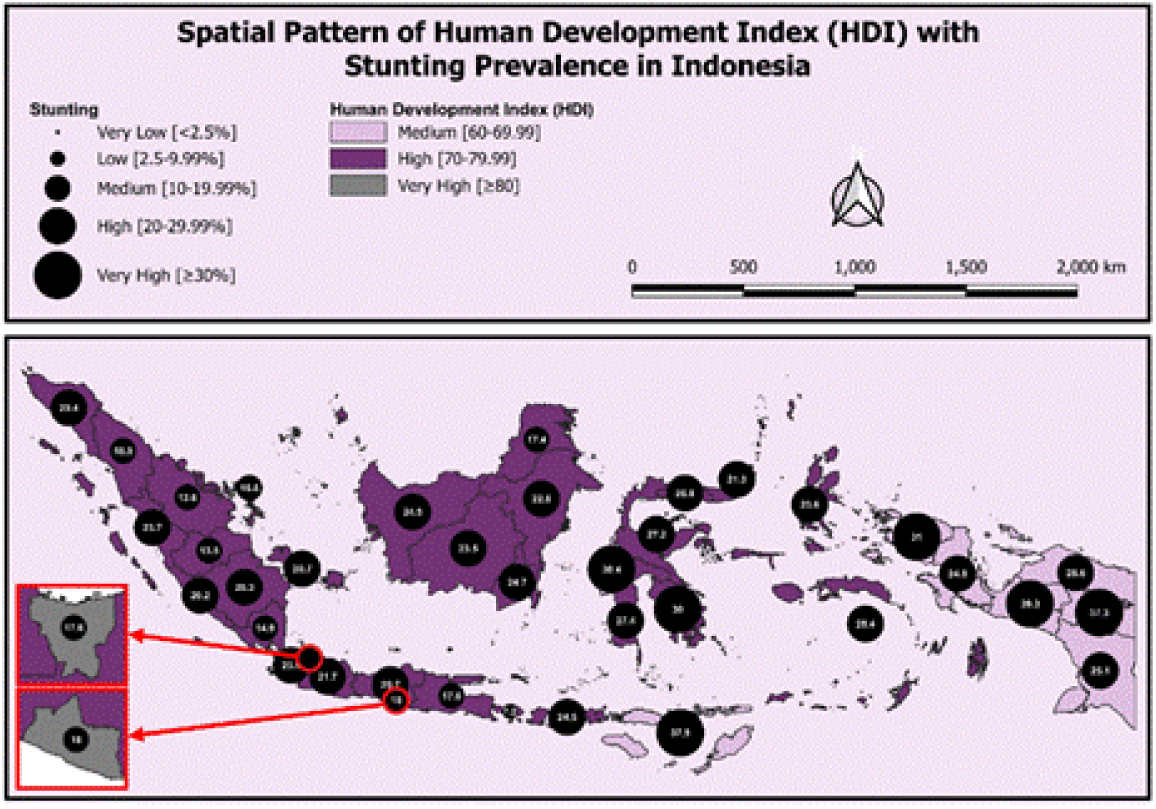
HDI pattern with Stunting Prevalence in Indonesia in 2023 Figure 1 illustrates the spatial relationship between HDI and stunting prevalence. In our findings, several provinces, including Central Papua, East Nusa Tenggara, Mountains Papua, South-West Papua, West Sulawesi, and Southeast Sulawesi were observed to have a very high stunting prevalence (mean: 34.3%) and were categorized as Medium-High HDI category. However, Bali, which is categorized as High HDI has the lowest stunting rate at 7.2%.

**Fig 2.**
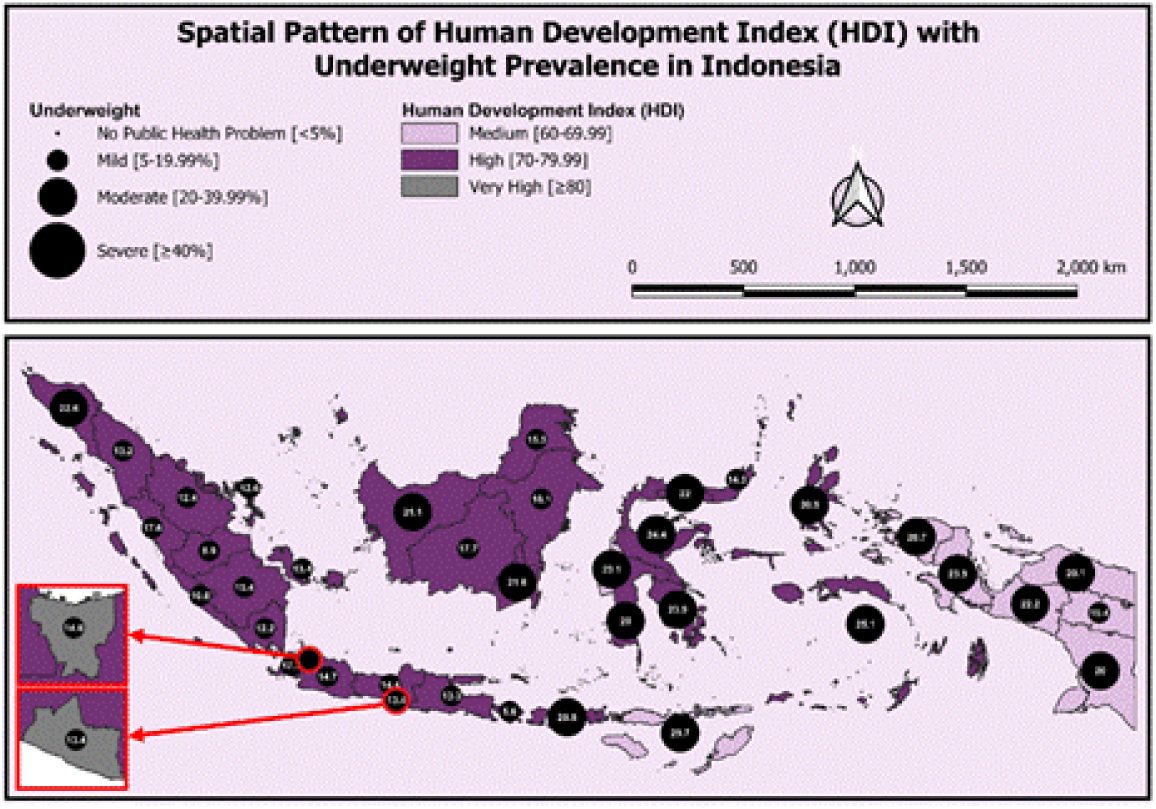
HDI pattern with Underweight Prevalence in Indonesia in 2023 Figure 2 illustrates the spatial relationship between HDI and underweight prevalence. Our analysis shows that East Nusa Tenggara has the highest prevalence in underweight at 29,7% and was categorized in Medium HDI. In contrast, Bali, which is categorized as High HDI province, recorded the lowest underweight prevalence at 5.6%.

**Fig 3.**
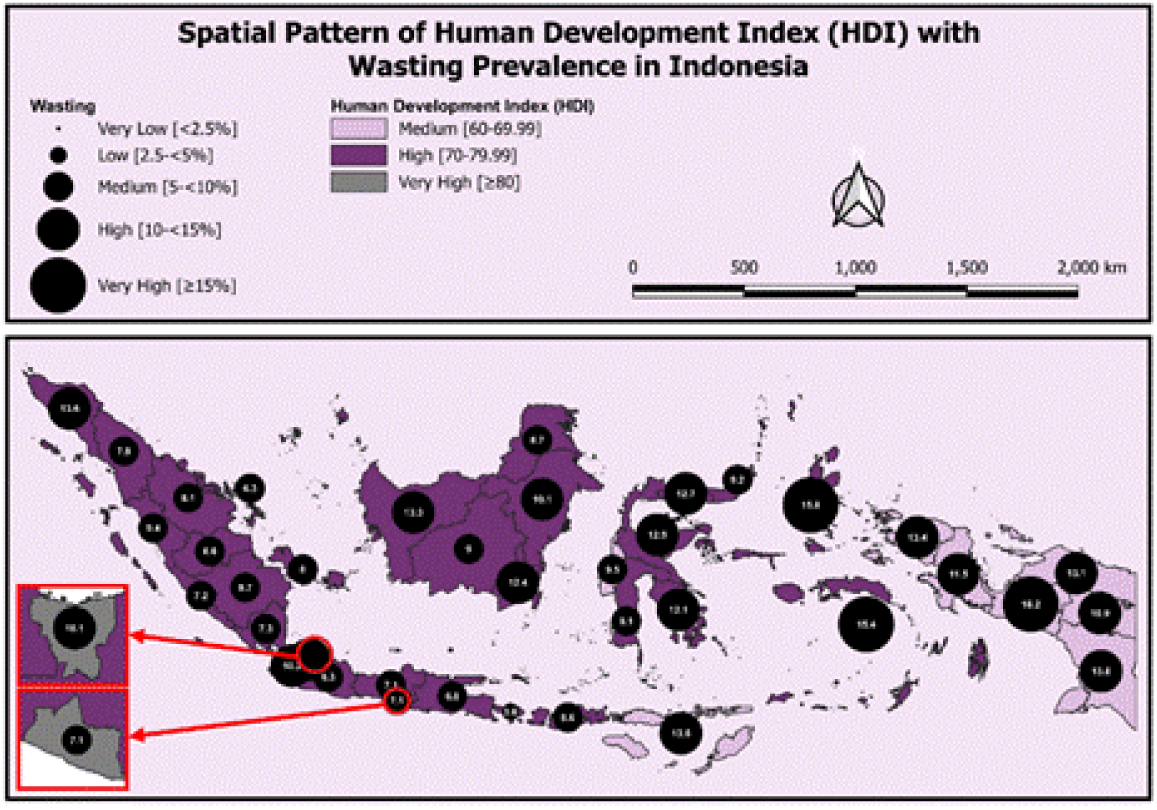
HDI pattern with Wasting Prevalence in Indonesia in 2023 Figure 3 illustrates the spatial relations between HDI and wasting prevalence. In our analysis, we found that Central Papua that categorized into Medium HDI has mild prevalence of wasting at 18,2%. In the other hand, Bali was observed as high has the lowest prevalence of wasting at 3,6%.

**Fig 4.**
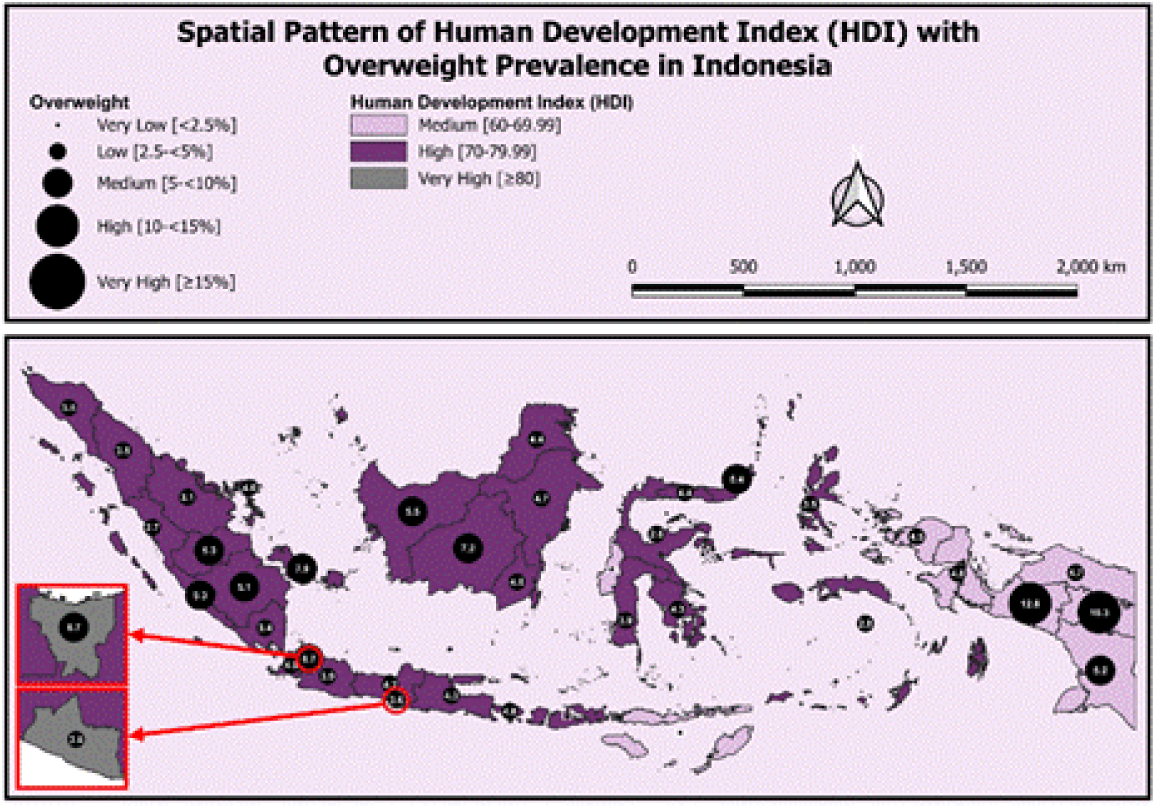
HDI pattern with Overweight Prevalence in Indonesia in 2023 Figure 4 illustrates the spatial relations between HDI and overweight prevalence. Our analysis found that Central Papua and Highlands Papua has the highest prevalence of overweight (mean: 11,6%), which categorized as Medium HDI province. While West Nusa Tenggara reported that it has very low prevalence of overweight at 1,7%, which are categorized as High HDI provinces. We also observe an increase in overweight prevalence as the HDI rises in a province.

According to the spatial analysis between HDI and nutritional status showed the prevalence of stunting, underweight, wasting, even overweight was found higher in Medium HDI provinces. There is an indication where double burden malnutrition is more likely emerging in this area. Studies across multiple LMICs reveal high prevalence of both underweight and overweight among children under five.

### Malnutrition Prevalence Differences by HDI Categories

Table 2 highlights the disparities in malnutrition prevalence based on HDI categorizations. The findings indicated that provinces with lower HDI levels experience a greater burden of undernutrition. Stunting rates were notably higher in low-to-medium HDI provinces (31.80%) compared to high-to-very-high (21.36%). Similarly, underweight prevalence was elevated in lower-HDI areas (21.89%) versus higher-HDI areas (16.60%). Wasting follows the same trend, with a prevalence of 13.00% in low-to-medium HDI provinces, significantly exceeding the 9.49% observed in high-to-very-high HDI regions. Unexpectedly, overweight was slightly more prevalent in low-to-medium HDI provinces (5.90%) than in high-to-very-high HDI provinces (4.40%), suggesting that the double burden of malnutrition may be affecting lower-HDI regions.

**Table 1.** Descriptions of Variables.

| Variable | Description |
| --- | --- |
| **Dependent Variables** |  |
| Stunting | Prevalence of children under five with height-for-age Z-score below -2 standard deviations relative to the WHO's growth standards (16). |
| Underweight | Prevalence of children under five with weight-for-age Z-score below -2 standard deviations relative to the WHO's growth standards (16). |
| Wasting | Prevalence of children under five with weight-for-height Z-score below -2 standard deviations relative to the WHO's growth standards (16). |
| Overweight | Prevalence of children under five with weight-for-age Z-score above +2 standard deviations relative to the WHO's growth standards (16). |
| **Independent Variables** |  |
| Human Development Index (HDI) | Summary measure of average achievement in key dimensions of human development: a long and healthy life, being knowledgeable, and having a decent standard of living (2). |
| Low birth weight | Percentage of births with a reported birth weight less than 2.5 kilograms regardless of gestational age (17). |
| Exclusive breastfeeding | Percentage of infants aged 0-5 months who received only breast milk (17). |
| Proper infant and young child feeding practices | Proportion of children aged 6-23 months who receive a minimum acceptable diet (17). |
| Adequate child's micronutrient (vit A) intake | Percentage of children consuming foods rich in vitamin A and receiving vitamin A supplements (17). |
| Proper handwashing practices | Percentage of households in which the place for hand washing was observed (17). |
| Proper sanitation | Percentage of households with a private toilet (17). |
| Complete basic immunization | Percentage of children aged 12-23 months who received specific vaccines based on the MOH 2017 (17). |
| Complete compulsory education of parents | Percentage of the population has completed senior high school (17). |

**Table 2.** Prevalence of Malnutrition by Human Development Index (HDI) Categories (%)

| HDI | Stunting | Underweight | Wasting | Overweight |
| --- | --- | --- | --- | --- |
| Low-Medium | 31.80 | 21.89 | 13.00 | 5.90 |
| High-Very High | 21.36 | 16.60 | 9.49 | 4.40 |
| Mean Differences | 10.44* | 5.29* | 3.51* | 1.50 |

\*Statistical significant at $p < 0.05$ , Mann-Whitney U test.

### Determinants of Malnutrition

Table 3 presents the prevalence of stunting, underweight, wasting, and overweight and their correlations with several determinants or characteristics. The mean prevalence of stunting, underweight, and wasting was 23.56%, 17.71%, and 10.22%, respectively, while overweight prevalence remained lower at 4.72%. Several significant correlations were observed in this analysis.

**Table 3.**
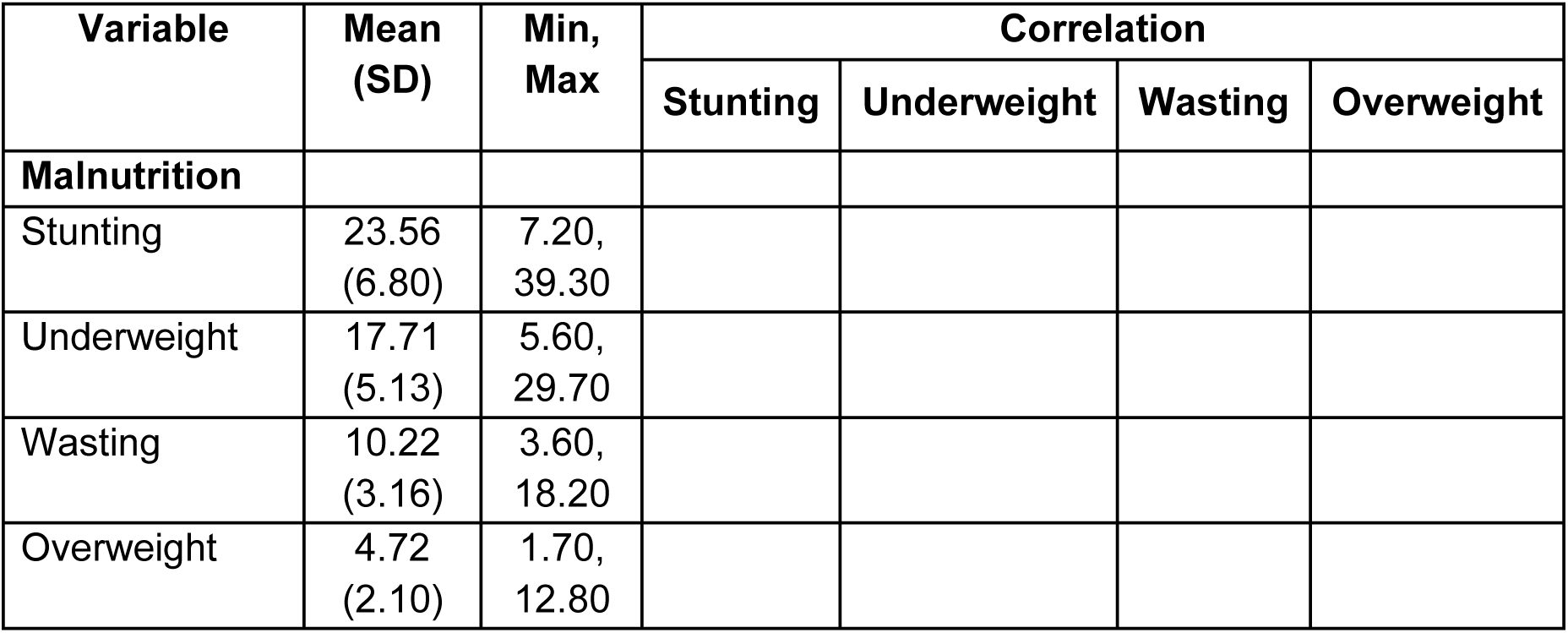
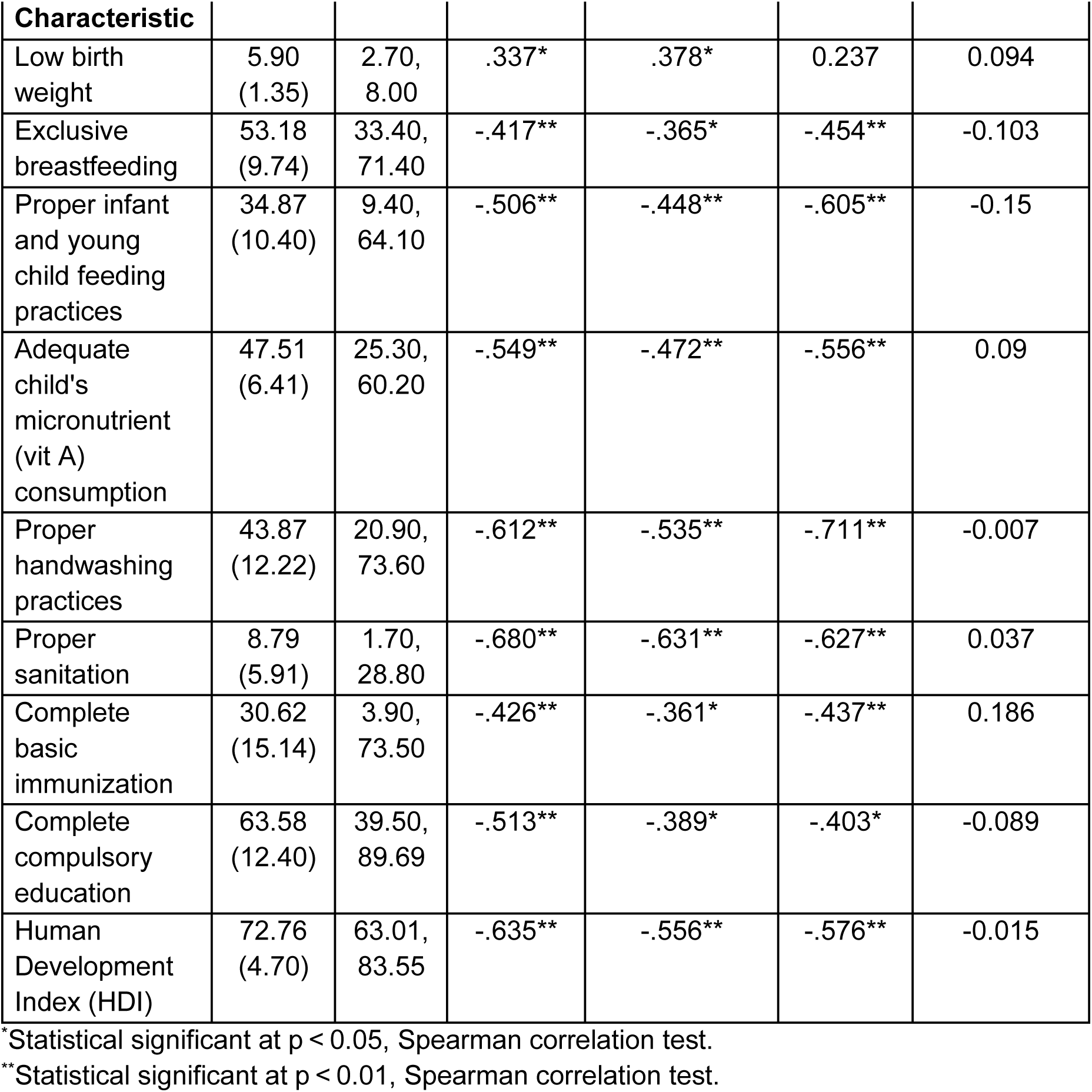
Prevalence of Malnutrition and Its Determinant.

Low birth weight showed a positive correlation with stunting (r=0.337, p < 0.05) and underweight (r = 0.378, p < 0.05). Exclusive breastfeeding exhibited significant negative correlations with stunting (r= −0.417, p<0.01), underweight (r=-0.365, p< 0.05), and wasting (r=-0.454, p< 0.01), indicating its protective effect against malnutrition. Similarly, proper infant and young child feeding practices were strongly associated with lower stunting (r=-0.506,p< 0.01), underweight (r=-0.448, p< 0.01), and wasting (r=-0.605, p<0.01). Nutritional adequacy was also a key determinant, as adequate vitamin A consumption correlated negatively with stunting (r=-0.549, p<0.01), underweight (r=-0.472, p<0.01), and wasting (r=-0.556, p< 0.01).

Proper handwashing practices and sanitation were among the strongest protective determinants against stunting, underweight, and wasting, with sanitation showing a particularly strong correlation with stunting (r=-0.680, p<0.01). Education and socioeconomic factors also played a significant role. Higher completion rates of basic education were significantly correlated with lower stunting (r=-0.513, p<0.01), underweight (r=-0.389, p<0.05), and wasting (r=-0.403, p<0.05). HDI was negatively correlated with stunting (r = −0.635, p < 0.01), underweight (r = −0.556, p < 0.01), and wasting (r = −0.576, p < 0.01). Interestingly, overweight prevalence did not show significant correlations with any determinants.

## Discussions

### Main Findings

This study aimed to examine difference HDI level impacts on nutritional outcomes and examine the determinants of malnutrition. In this paper we categorized undernutrition prevalence (stunting, underweight, and wasting) based on Low-Medium HDI and High-Very High HDI provincial data. Three main results can be drawn from this research: Firstly, we found 30 provinces, mostly from western part of Indonesia, indicated as High-Very High HDI provinces with significantly lower prevalence in undernutrition in all forms. Bali, for instance, consistently showing lowest rate at undernutrition prevalence. Second, the other 8 provinces, where mostly from central and eastern part of Indonesia indicated as Medium HDI areas with higher rates in all undernutrition forms. Third, exclusive breastfeeding, proper IYCF practice, child’s micronutrient consumption, proper handwashing, proper sanitation, complete basic immunization, and complete compulsory parent education significantly correlated with all forms of malnutrition (stunting, underweight, wasting) this finding proved that diets as immediate cause and sanitation as underlying cause contribute to child nutrition status as outcomes (7).

### Nutritional Status and HDI

The national prevalence of stunting, underweight, and wasting was significantly higher than the global percentage reported in 2017 (21,5%, 15,9%, and 8,5%), and the prevalence of overweight indicated lower than global report 4,2% (19,20). In 2022, Indonesia was reported as a High-HDI country along with the Philippines and Vietnam in Southeast Asia (21). The double burden of malnutrition (DBM), characterized by the coexistence of undernutrition and overweight/obesity. This phenomenon affects women of reproductive age and children, with prevalence varying across Southeast Asian countries (22). The previous research was in line witn our analysis, where we identified Medium HDI provinces with relatively high prevalence in undernutrition and a growing number of overweight.

The lower HDI implies the lower life expectancy, years of schooling, and income, which reflect the GNI index. In spite of our limitation to examine each dimension of HDI, an evidenced based demonstration shows that GNI, one of the components of HDI linked with undernutrition form. Like poverty, undernutrition is a cycle that occurs in an intergenerational cycle, contributing to food insecurities, infections, and allostatic load (the body’s accumulated wear and tear as a result of adjusting to negative physical or psychological stress) (23). Despite decades of nutritional interventions, early childhood growth failure in low-income countries (LMICs) persists. Children from low-income families experience poorer growth, neurocognitive outcomes, and educational attainment, leading to lower economic productivity and intergenerational transmission of poverty and undernutrition (24).

Lower HDI is also associated with higher rates of infectious disease (25). Infections induce a significant nutritional demand on the host, affecting both metabolism and immune function and acute infections can lead to long-term alterations in the gut microbiota, promoting host adaptation to nutrient restriction (26), by which an immediate cause of undernutrition. In other mechanisms, infections affect nutritional demand on biological systems through inflammation, which triggers anorexia and reduced food intake (27). Furthermore, undernutrition and infection form a vicious cycle. Malnutrition compromises the immune system and leads to increased susceptibility to infections (28) while infections worsen nutritional status through mechanisms like nutrient malabsorption that leads to severe undernutrition.

Inadequate nutrition intake, poor water, sanitation, and hygiene (WASH) practice, and inequality of access to health services are some key factors influencing high prevalence in stunting (29). However, since 2015 Indonesian government has implemented village fund programs, but the challenges remains on its effectiveness and program implementation (30). In 2018, the government started the implementation of stunting convergence action in selected 100 districts/cities across Indonesia with the highest number of stunting prevalence (31). Furthermore, addressing stunting requires a multisectoral approach, integrating nutrition-specific and nutrition-sensitive interventions, while focusing on improving WASH programs and health services especially in rural and remote areas (32,33).

Undernutrition has been the most prevalent type of malnutrition in low- and middle-income countries (LMICs). However, with ongoing economic development, urbanization, and shifts in population dynamics, overweight and obesity are becoming increasingly common, resulting in a double burden of malnutrition as we noticed in several provinces in this research (34).

As we indicates the rise of double burden malnutrition in Medium HDI provinces, this is a growing challenge in LMICs (35) .The nutrition transition, characterized by declining rates of undernutrition and increasing prevalence of overweight and obesity, is closely linked to economic development and urbanization in low- and middle-income countries (36). This phenomenon is driven by factors such as increased energy intake, overconsumption of processed foods, and reduced physical activity (37). hile the shift towards ultra-processed food consumption is widespread, it is not inevitable, and targeted policies can help mitigate its negative impacts (38).

### Determinants of Stunting, Wasting, Underweight, and Overweight

The findings suggest that various characteristics were associated with nutritional status. The Spearman correlation analysis showed a moderately strong and significant positive correlation between low-birth weight prevalence and stunting (r = 0.392) as well as underweight (r = 0.413), indicating that provinces with higher low birth weight prevalence tend to have higher rates of stunting and underweight. This finding is supported by a study in India that found children with low birth weight have a significantly higher risk of experiencing stunting compared to those with normal birth weight (39). Moreover, a meta-analysis investigated that children under five with a history of low birth weight were 2.21 times more likely to be underweight compared to those with a normal birth weight history (40). Low birth weight is a significant risk factor for stunting and underweight due to its long-term effects on growth and health. Infants with low birth weight often experience intrauterine growth restriction (IUGR), leading to inadequate nutrient stores and impaired organ development, which persist into early childhood (41). On the other hand, low birth weight conditions weaken the immune system, increasing susceptibility to infections, while infections further exacerbate malnutrition by impairing appetite, absorption, and metabolism (42).

A significant negative correlation was observed among the prevalence of exclusive breastfeeding and stunting, underweight, and wasting. The finding indicated that higher rate of exclusive breastfeeding was correlated with lower stunting, underweight, and wasting prevalence. This finding aligns with previous research underscoring the protective effects of exclusive breastfeeding against various forms of undernutrition. A systematic review and meta-analysis encompassing studies from developing countries found that exclusive breastfeeding is associated with a reduced risk of overall undernutrition (OR=0.82; 95% CI:0.68–0.99) and specifically stunting (OR=0.73; 95%CI:0.55–0.95) (43). Similarly, a study conducted in Eastern Indonesia reported that exclusively breastfed children aged 6–24 months had a lower likelihood of being stunted compared to their non-exclusively breastfed counterparts, even after adjusting for socioeconomic factors (44). These correlations can be attributed to the nutritional adequacy of breast milk, which provides essential nutrients and antibodies crucial for optimal growth and immune function during the first six months of life (45). Furthermore, exclusive breastfeeding supports healthy weight gain and linear growth, contributing to lower rates of underweight and wasting (46).

Proper infant and young child feeding (IYCF) practices were negatively correlated with stunting, underweight, and wasting. These findings align with a previous study demonstrating that adherence to recommended feeding guidelines is associated with lower rates of various forms of undernutrition among children aged 6 to 23 months (47). Such practices help to ensure adequate nutrient intake during the critical first two years of life, thereby supporting optimal growth and immune development. A key component of IYCF is ensuring minimum dietary diversity (MDD) during complementary feeding. Evidence has shown that children who achieve adequate MDD had lower odds of stunting, underweight, and wasting (48). Although earlier literature has reported a potential link between certain feeding practices and increased risk of overweight (49,50), particularly through excessive caloric intake or poor portion control, our analysis did not find a statistically significant correlation. This suggests that in the observed population, recommended feeding behaviors may primarily influence undernutrition, with overweight potentially shaped more by other contextual or behavioral factors not captured within the IYCF indicators.

This study finds a strong, moderate and negative correlation between adequate child’s vitamin A consumption and stunting, underweight, and wasting (r= −0.549, −0.472 and −0.556) illustrating provinces with lower consumed foods rich in vitamin A and children received vitamin A supplementation (VAS) tend to have higher rates in stunting, underweight, and wasting. Recent study confirms that vitamin A deficiency (VAD) is associated with stunting in Indonesian children aged 6-59 months (51) and the children who consumed vitamin-A and vaccination had a lower likelihood of wasting than their counterparts (52). Biological mechanism explains that VAD is linked to child growth failure due to a causal relationship with diarrheal diseases, as disrupted mucosal integrity reduces nutrient absorption due to the absence of vitamin A in gastrointestinal tract (53). Leading to decreased nutritional absorptions, increased vulnerability to infectious diseases and malnutrition which result in growth failure (53,54). Several studies have shown that food fortification has been effective in reducing vitamin A deficiency in infants and children (55,56).

On the other hand, there are remaining challenges influencing VAS coverage include maternal education, household income, media exposure, antenatal/postnatal care utilization, and regional disparities (57,58). Thus, interventions should be focused to increase awareness through media campaigns, promoting antenatal and postnatal care, rural residents, and lower-income households to be encouraged to practice regular consumption of foods high in vitamin A and access VAS since it plays a crucial role in child health, immunity, and growth.

The findings of this study revealed a strong negative correlation between proper handwashing practices and the prevalence of stunting (r=-0.612), underweight (r=-0.535), and wasting (r=-0.711), suggesting that the greater the coverage of proper handwashing, the lower the rates of those malnutrition. Similar results were reported in Bangladesh, where children under five in homes with limited handwashing facilities were more prone to stunting than their peers (59) as well as in Benin, West Africa, which found a 1.33 times higher risk of underweight in children under five without access to basic hygiene (60). Additionally, a study in Nepal found that increased handwashing was positively and significantly associated with weight-for-height during the early months of the study among Nepalese children (61). Underlying biological mechanisms include increased susceptibility to intestinal infections due to water and surface contamination, which further impairs nutrient absorption and decreases nutritional status to the point of worsening body mass index (62). These mechanisms reinforce the evidence that sanitation interventions, including handwashing promotion, play a role in improving nutritional status and body weight.

Furthermore, there is a strong negative correlation between adequate sanitation coverage and stunting (r=-0.680), underweight (r=-0.631), and wasting (r=-0.627), i.e., the higher the access to sanitation, the lower the prevalence of undernutrition. This finding is supported by a study in Southern Punjab, Pakistan, which confirmed that poor sanitation conditions are a major contributing factor to stunting in children under five years old (63). In Ethiopia, the absence of household toilets was associated with a significantly increased risk of underweight (64). In addition, each percentage increase in sanitation facilities in India decreased the prevalence of wasting by 0.1-0.3% (65). Fecal contamination in the home environment, due to poor sanitation and not washing hands after defecation, facilitates the spread of environmental enteropathies that damage gut integrity and inhibit nutrient absorption (66).

Collectively, poor WASH (water, sanitation, and hygiene) factors mediate malnutrition through three main pathways: (1) recurrent diarrhea that stunts growth; (2) earthworm infection, particularly during pregnancy, which triggers maternal malabsorption and anemia resulting in neonatal stunting; and (3) environmental enteric dysfunction (EED), where chronic inflammation of the gut weakens barrier function and nutrient absorption (67). These three mechanisms emphasize the crucial role of sanitation interventions, including handwashing promotion, in breaking the cycle of pathogen transmission, reducing chronic inflammation, and improving the nutritional status of children under five (68).

Our study finds that complete basic immunization has moderately strong and negative correlation with stunting, underweight and wasting (r= −0.426, −0.404 and −0.535), which implies the lower coverage of complete basic immunization in provinces tend to have higher prevalence of underweight and wasting. In 2023, only 35.8% of children aged 12–23 months in Indonesia received complete vaccination, with coverage as low as 4.0% in Highlands Papua—highlighting significant barriers and disparities in program implementation (14). The coverage rates are far from WHO and UNICEF’s global immunization goals, which outline a goal of 80% coverage (69). Our findings are in line with the low coverage immunization in India that influences nutritional status (70). Another study conducted in several countries showed that incomplete vaccination leads to underweight and wasting among children under-five (71,72). Conversely, complete vaccination schedules are associated with better nutritional outcomes in children (70). In our understanding, children with incomplete vaccination are likely to be exposed to infection and disease that leads to malnutrition, but at the same time, malnourished children could impaired vaccine efficacy and increase their susceptibility to disease like polio and rotavirus (73). Despite our failure to see each coverage immunization status, it is important to monitor the coverage for vaccines that require multiple doses. Compared to children in areas with the highest healthcare facilities, children in areas with the fewest public health facilities typically receive less comprehensive immunization coverage (74). To address this challenges, improving healthcare access, including strengthening vaccination programs in communities by education campaigns as a nutrition-sensitive intervention during the first 1000 days of life is crucial to increase immunization coverage and reduce childhood morbidity and mortality from vaccine-preventable diseases.

Our analysis finds a negative correlation and moderately strong correlation between complete compulsory education with stunting (r= −0.513) underweight (r= −0.453), and wasting (r= −0.403), indicating provinces with lower rates of parents completing senior high school tend to have higher stunting and underweight prevalence. This finding consistent with several systematic review studies which conclude that parental education consistently emerges as a crucial factor influencing child stunting, with lower education levels associated with higher stunting risks (75,76). Households with higher parental education were more likely to experienced double burden malnutrition, including in undernutrition form (77). The possible explanation for this causal is higher average education in a community is associated with improved individual health outcomes and health-seeking behaviors (78). Lower levels of education are associated with less dietary diversity, less healthful eating habits and likely unaware of proper dietary feeding practice (79).

Our analysis found negative correlation among HDI and the prevalence of stunting, underweight, and wasting. Our finding corroborates earlier research indicating that improvements in socio-economic development, particularly in women’s education, household income, and access to healthcare, are associated with reductions in child undernutrition (stunting, underweight, wasting). For instance, analyses from DHS Kenya and pooled LMIC data identified socioeconomic status and parental nutrition as the primary determinants of undernutrition (80). In Indonesia, results from the Indonesia Family Life Survey (IFLS, 2014) showed that maternal education and household expenditure explain over 55% of the stunting disparities across socio-economic groups (81). Moreover, the significant association between HDI and wasting, which is often considered an acute form of malnutrition, suggests that broader development may also enhance community resilience to short-term shocks such as infections or food shortages, which often precipitate wasting (82). In contrast, we did not find a statistically significant correlation between HDI and overweight. This differs from some previous studies that have reported a positive association between HDI and overweight or obesity, especially in contexts undergoing rapid nutrition transition (38,83,84). The absence of such a correlation in our analysis may reflect the complex and multifactorial nature of overweight, which is not solely driven by socio-economic development, but also influenced by lifestyle behaviors, food environments, and cultural norms that vary across regions regardless of HDI level.

This study has several strengths and limitations. To our knowledge, our study provides the first analysis of the intersection between HDI and the determinants of malnutrition in Indonesia, including the use of spatial analysis to map HDI and nutritional status across all provinces in Indonesia. This approach provides a comprehensive visualization of regional disparities on nutritional outcomes. Additionally, the study utilizes a large, nationally representative dataset, enhancing the generalizability of the findings. However, some limitations should be acknowledged. The cross-sectional design prevents causal inferences between HDI and nutritional outcomes. Moreover, while HDI captures broad socio-economic factors, it may not fully account for localized determinants such as food security, healthcare accessibility, and cultural dietary practices.

Future research should explore the causal mechanisms underlying the relationship between HDI and nutritional status by utilizing longitudinal data. Investigating how specific HDI components, such as education or healthcare access, independently influence nutritional outcomes could provide deeper insights for targeted interventions. Additionally, studies incorporating more granular, district-level data and contextual factors like food security, dietary patterns, and healthcare infrastructure would help refine the understanding of regional disparities. Given the double burden of malnutrition observed in different HDI categories, future research should also examine the effectiveness of integrated policies addressing both undernutrition and overnutrition within the same populations.

## Conclusions

This study aimed to examine the relationship between the Human Development Index (HDI) and nutritional status in Indonesia, utilizing spatial analysis to highlight disparities across provinces. Our findings reveal a clear trend that provinces with lower HDI experience significantly higher rates of undernutrition, including stunting, underweight, and wasting. This finding underscores the complex interplay between socio-economic development and nutritional outcomes. Furthermore, factors such as exclusive breastfeeding, proper IYCF practice, child’s micronutrient consumption, proper handwashing, proper sanitation, complete basic immunization, and complete compulsory parent education, emerged as critical determinants influencing undernutrition prevalence. These insights have practical implications for public health policies and interventions. Addressing undernutrition in lower HDI regions requires a multi-sectoral approach, integrating improvements in education, sanitation, and healthcare access. Future studies should incorporate longitudinal data to better understand the dynamic relationship between HDI and nutritional outcomes. Additionally, exploring localized determinants, such as food security and healthcare infrastructure at the district level, could refine intervention strategies. In conclusion, our findings emphasize the urgent need for region-specific public health interventions to tackle malnutrition problems in Indonesia.

## Data Availability

All data produced in the present study are available upon reasonable request to the authors

